# Sex-based clinical and immunological differences in COVID-19

**DOI:** 10.1101/2020.08.29.20126201

**Authors:** Kening Li, Bin Huang, Yun Cai, Zhihua Wang, Lu Li, Lingxiang Wu, Mengyan Zhu, Jie Li, Ziyu Wang, Min Wu, Wanlin Li, Wei Wu, Lishen Zhang, Xinyi Xia, Shukui Wang, Qianghu Wang

**Author notes:** Kening Li, Bin Huang, Yun Cai, and Zhihua Wang contributed equally to this article. **Correspondence to:** Xinyi Xia,; Shukui Wang,; Qianghu Wang.

## Abstract

**Background:** Males and females differ in their immunological responses to foreign pathogens. However, most of the current COVID-19 clinical practices and trials do not take sex as consideration.

**Methods:** We performed an unbiased sex-based comparative analysis for the clinical outcomes, peripheral immune cells, and SARS-CoV-2 specific antibody levels of 1,558 males and 1,499 females COVID-19 patients from a single center. The lymphocyte subgroups were measured by Flow cytometry. Total antibody, Spike protein (S)-, receptor binding domain (RBD)-, and nucleoprotein (N)-specific IgM and IgG levels were measured by chemiluminescence.

**Results:** We found that the mortality and ICU admission rates were approximately 2-fold higher in males than that in females (P<0.005). Survival analysis revealed that sex is an independent prognostic factor for COVID-19 (Hazard ratio=2.2, P=0.003). The concentration of inflammatory factors in peripheral blood was significantly higher in males. Besides, the renal and hepatic abnormality induced by COVID-19 was more common in males during the hospitalization. The analysis of lymphocyte subsets revealed that the percentage of CD19+ B cell and CD4+ T cell was significantly higher in females (P<0.001) during hospitalization, indicating the stronger humoral immunity in females than males. Notably, the protective IgG sharply increased and reached a peak in the fourth week after symptom onset in females, while gradually increased and reached a peak in the seventh week in males.

**Conclusions:** The unfavorable prognosis of male COVID-19 patients may result from the weak humoral immunity and indolent antibody responses during SARS-CoV-2 infection and recovery. Early medical intervention and close monitoring are important, especially for male COVID-19 patients. Hormonal or convalescent plasma therapy may help improve the immunity of males to fight against SARS-CoV-2 infection.

## Background

Caused by the infection of severe acute respiratory syndrome coronavirus 2 (SARS-CoV-2), The outbreak of novel coronavirus disease 2019 (COVID-19) is a worldwide pandemic spreading in more than 210 countries and territories[1, 2]. As of May 8, 2020, a total of 3,672,238 confirmed cases were reported, of which 254,045 patients died (WHO situation report 108). Approximately 100,000 confirmed cases increased every single day, extremely challenging the public health and medical service around the globe. Investigating the risk factors of susceptibility and prognosis for COVID-19 is necessary to help disease prevention and precise therapy.

According to the previous reports, age is a risk factor of COVID-19[3]. In a report of 1099 patients with COVID-19 from 552 hospitals in 30 provinces in China, patients with severe disease were older than those with the non-severe disease by a median of 7 years[4]. As the fact that SARS-CoV-2 has an approximately 85% nucleotide sequence identity to SARS-CoV[5], which was the causal agent of the severe acute respiratory syndrome outbreaks in 2003, the epidemiological risk factors may be similar between SARS-CoV and SARS-CoV-2. In addition to age, epidemiological studies showed that the incidence and mortality of SARS-CoV infection were sex-dependent[6]. Males were more susceptible, and experienced more severe disease after SARS-CoV infection [7]. A recent case series study reported that 75% of patients who died of COVID-19 were men[8]. Moreover, some researchers proposed that clinical trials for COVID-19 should include sex as a variable because of the biological difference between males and females[9]. Notably, experiments in mice indicated that ovariectomy or treating female mice with an estrogen receptor antagonist increased mortality after SARS-CoV infection [10], suggesting the hormonal effect plays an important role in the immune response against infection. However, the sex-based clinical outcome, as well as the underlying biological difference in COVID-19 is still unclear. In this study, by describing the clinical and laboratory characteristics of 3,057 COVID-19 patients from a single center, we performed an unbiased sex-based comparative analysis for the clinical, cellular and molecular differences in COVID-19. Our results will provide important information for the epidemiology and precise therapy for this emergent pandemic.

## Methods

### Patients

We analyzed the laboratory test results of 3,057 COVID-19 patients, including 1,455 mild or moderate, 1,417 severe, and 150 critical cases, admitted from February 4 to March 30 at Wuhan Huoshenshan Hospital. The severity degree of each patient was determined according to the clinical classification criterion in Diagnosis and Treatment Protocol for Novel Coronavirus Pneumonia released by the National Health Commission (trail version 7). We obtained the clinical characteristics and laboratory findings of all patients from the electronic medical records of the hospital. This study was approved by the Medical Ethical Committee of Wuhan Huoshenshan Hospital. Written informed consent was obtained from each patient.

### The lymphocyte subgroup assay

The lymphocyte subgroups were measured by Flow cytometry (CytoFLEX flow cytometry system, Beckman coulter, Inc.) using commercially available kits (Beckman coulter, Inc.) according to the manufacture’s protocol. Briefly, the reagents of the BD six-color lymphocyte subgroup (FITC-CD3, PE-CD16/PE-CD56, PerCP-Cy5.5-CD45, PE-Cy7-CD4, APC-CD19, and APC-Cy7-CD8) were mixed with the whole blood and incubated at room temperature for 20 minutes, followed by adding 1mL of a lysis solution with 30 minutes incubating. The proportion of CD3+, CD3+/CD4+, CD3+/CD8+, CD3-/CD19+, CD3-/CD56+/CD16+ cells in lymphocytes was analyzed with the software.

### Serum anti-SARS-CoV-2 antibodies assay

Total SARS-CoV-2 IgM or IgG in the serum was measured by chemiluminescence using commercially available kits (Shenzhen YHLO Biotech Co., Ltd.), which was coated with N and S proteins, in 1850 patients at different time points. 416 of these patients were tested for S-specific, RBD-specific, and N-specific IgM and IgG levels at different time points by chemiluminescence using commercially available kits (Nanjing RealMind Biotech Co., Ltd.), including 126 mild or moderate patients, and 290 severe or critical patients. Briefly, the blood samples were centrifuged at room temperature, the supernatant was taken and incubated with antigen-coated magnetic beads. The antigen-antibody complex is then captured, incubated, and reacted with hydrogen peroxide in an excitatory buffer. Relative luminescence intensity was recorded in the ACL2800 chemiluminescence system (Nanjing RealMind Biotech Co., Ltd.). The relative luminescence intensity was converted to AU/ML antibody levels.

### Definition of physiological abnormalities

We identified the cardiac abnormal patients based on the level of B-type natriuretic peptide (BNP). BNP levels greater than the maximum of the normal range were considered as an abnormality. The abnormality of the level of creatinine (CRE) and/or blood urea nitrogen (BUN) was used to recognize the patients with renal abnormality. Besides, patients who had glutamic-pyruvic transaminase (ALT), glutamic oxalacetic transaminase (AST), alkaline phosphatase (ALP), and/or glutamyltranspeptidase (GGT) 2-fold greater than the normal upper limit were thought to be hepatic abnormal.

### Survival analysis

Survival was estimated according to the Kaplan–Meier method. The log-rank test was used to assess statistical significance. To recognize the risk factors for COVID-19, age, sex, pre-existing diseases, days from symptoms onset to admission, and days from admission to discharge were evaluated in univariable Cox regression models for outcome. P values < 0.05 were considered statistically significant. The significant factors of univariable analysis were further analyzed by multivariable Cox proportional hazards model.

### Statistical analysis

We used the Wilcoxon rank-sum test or Fisher’s exact test to compare the difference between groups where appropriate. Continuous and categorical variables were presented as median (IQR) and n (%), respectively.

## Results

### Sex is an independent prognostic factor for COVID-19

To evaluate the relationship between sex and COVID-19 susceptibility and prognosis, we compared the clinical characteristics and outcomes between male and female patients (Table 1). There were 1,558 males and 1,499 females in our cohort, implying that the susceptibility to SARS-CoV-2 might be not associated with sex. Patients with the pre-existing chronic obstructive pulmonary disease in males were more than in females (96 (6.2%) vs. 51 (3.4%) in males and females, respectively, *P*<0.001), possibly because of the much higher smoking rate in men than in women in China (288 million men vs 12.6 million women were smokers in 2018) [11]. Besides, pre-existing chronic liver disease was more common in males (57 (3.7%) vs. 26 (1.7%) in males and females, respectively, *P*=0.002). Although the hospitalization time had no significant difference between sexes, the severity of COVID-19 was significantly associated with sex (*P*=0.002), as the percentages of critically ill patients were 6.2% vs. 3.5% in males and females, respectively. During the hospitalization, 73 (4.69%) males and 41 (2.74%) females got admitted to the ICU eventually (*P*=0.005). Notably, the mortality was more than 2-fold higher in males than that in females, with 46 (3.0%) males and 21 (1.40%) females died (*P*=0.004) in our cohort (Table 1). To further assess the outcomes of different sexes, we performed survival analysis for 3,057 COVID-19 patients (Figure 1A). Results showed that males had significantly unfavorable outcomes (log-rank test, *P*=0.003, HR=2.1, 95% IC: 1.3-3.6). By integrating age, sex, hospitalization time, and various pre-existing diseases to perform univariable and multivariable Cox Regression (Table 2, Supplementary Table S1), we found that sex was an independent risk factor for COVID-19 (H*P*=2.2, *P*=0.003, 95% IC: 1.31-3.74). Besides, age and intervals from symptoms onset to admission were also significant independent risk factors for COVID-19, indicating the crucial role of timely medical intervention for COVID-19 patients.

**Table 1.**
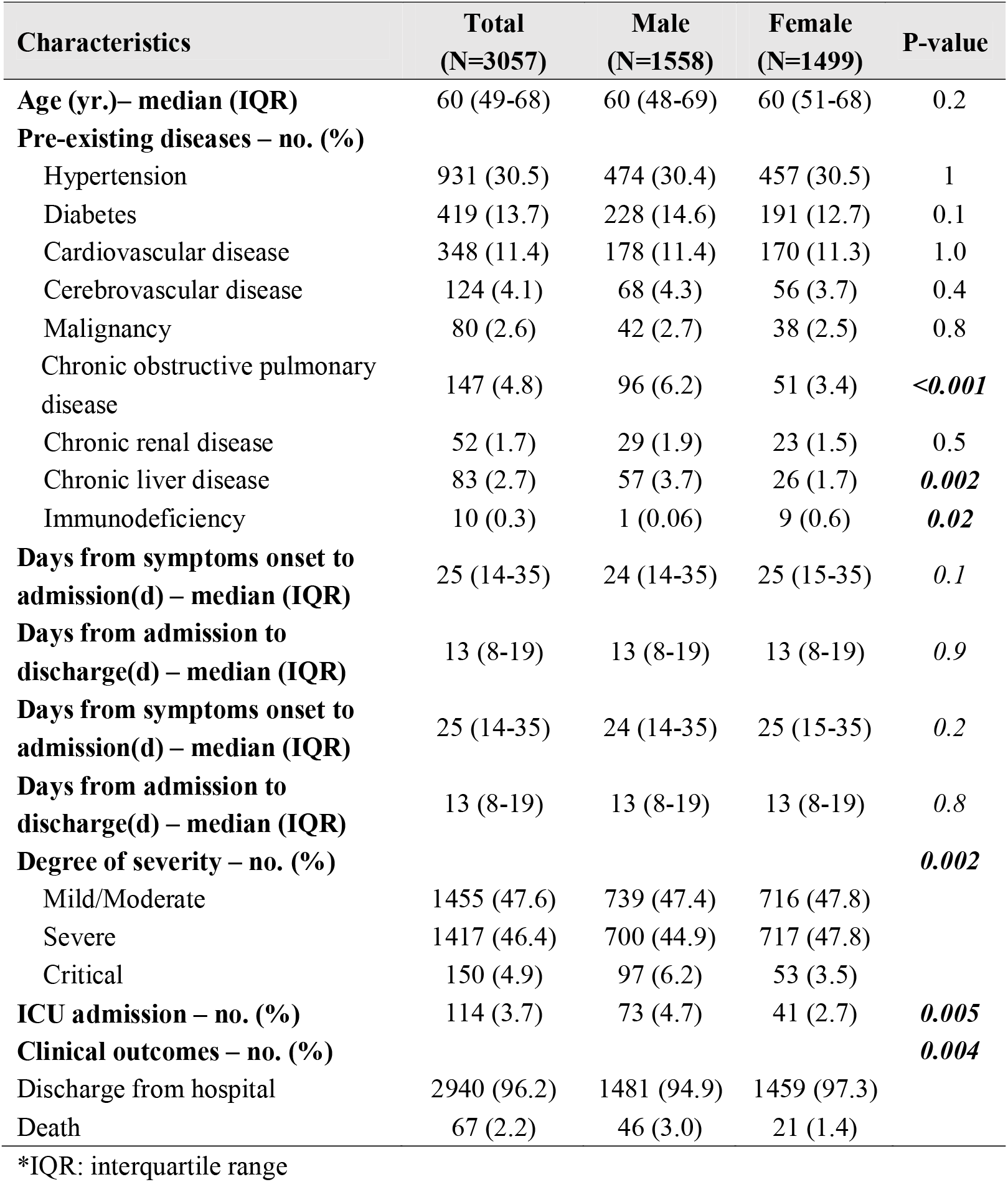
Comparison of Clinical Characteristics and outcomes between Males and Females

**Figure 1.**
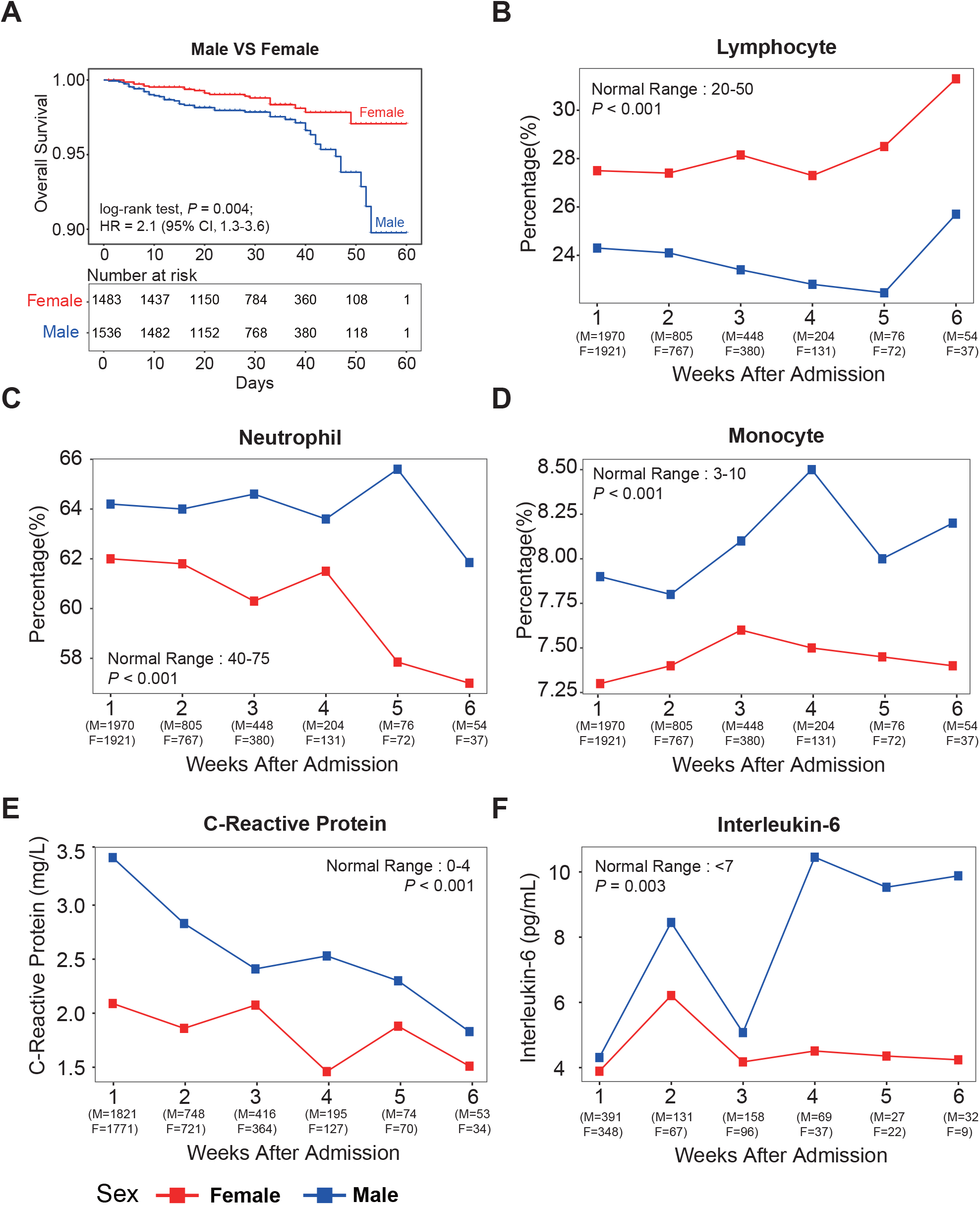
Survival analysis and comparison of prognostic indicators between males and females. (A) Sex-based survival analysis of COVID-19 patients. (B-E) Dynamic changes of laboratory findings in male and female patients. The x-axis displays the weeks after admission. F indicates the number of tests of females, and M indicates the number of tests of males. The y-axis displays the level of prognostic indicators. Red line based on median is used to profile the variation tendency of the females, and blue line based on median is used to profile the variation tendency of the males.

**Table 2.** Multivariable analyses of survival of COVID-19 patients

| Variable | HR<sup>*</sup> | 95% CI<sup>*</sup> | P-value |
| --- | --- | --- | --- |
| Age | 1.1 | 1.0-1.1 | *<0.001* |
| Sex | 2.2 | 1.3-3.7 | *0.003* |
| Days from symptoms onset to admission(d) | 0.9 | 0.9-1.0 | *<0.001* |

<sup>\*</sup>HR: hazard ratio, 95%CI: 95% confidence interval

Furthermore, we compared the levels and dynamic changes of laboratory indicators associated with COVID-19 prognosis between different sexes. Previous studies reported that the ratio between neutrophil and lymphocyte percentage is an important index for the prognosis of COVID-19 [12]. Patients with lower lymphocyte percentage and higher neutrophil percentage often have a poor outcome[13]. Our results showed that the lymphocyte percentage was significantly lower in males than that in females throughout the hospitalization (23.0 [14.2-30.4] % vs. 27.0 [19.2-33.5] % in males and females, respectively, *P*<.001, Figure 1B). On the contrary, neutrophil (65.4 [57.1-75.7] % vs. 62.4 [55.2-70.0] % in males and females, respectively, *P*<0.001, Figure 1C) and monocyte percentages (7.7 [6.2-9.3] % vs. 7.2 [5.9-8.7] % in males and females, respectively, *P*<0.001, Figure 1D) were higher in males than that in females throughout the hospitalization. Besides, the concentration of C-reactive protein (CRP) was significantly higher in males during hospitalization (3.5 [1.1-17.3] mg/L vs. 2.1 [0.8-7.4] mg/L in males and females, respectively, *P*<0.001), suggesting the increased inflammatory responses in males (Figure 1E). Meanwhile, the level of inflammatory cytokine Interleukin-6 (IL-6) was higher in males (5.9 [2.9-30.9] pg/mL vs. 4.4 [2.6-15.3] pg/mL in males and females, respectively, *P*=0.003), especially that the median IL-6 level in males was significantly higher than the normal range, while IL-6 in females maintained a relatively low level after 3 weeks of admission (Figure 1F). Recent studies reported that the elevated level of cytokines (or even cytokine storm) could lead to acute pulmonary injury and acute respiratory distress syndrome (ARDS), related to the ICU admission and death[14]. In our cohort, 12.0% (90/752) males had IL-6 concentration more than 5-fold of the upper limit of the normal range, while 7.1% (51/720) females reached this concentration during hospitalization (*P*=0.002). Thus, our results showed that males with COVID-19 were more prone to experience severe inflammation and increased cytokine level. In addition, we compared the number of patients with cardiac, renal, and hepatic function abnormality induced by COVID-19 between males and females during hospitalization (Supplementary Figure S1). Results showed that the percentage of COVID-19-induced renal (6.5% [102/1588] vs. 4.2% [63/1499] in males and females, respectively, *P*=0.002) and hepatic abnormalities were significantly higher in males than that in females (40.9% [650/1588] vs. 31.7% [475/1499] in males and females, respectively, *P*=0.003). It is important to closely monitor the cytokine levels and other indicators for male COVID-19 patients, and provide timely treatment.

### Males have weak humoral immunity during the infection and recovery of COVID-19

The latest study demonstrated that the humoral and cellular immune response plays an important role in defending against SARS-CoV-2 infection [15]. To explore the underlying immunological basis of the relatively poor clinical outcomes in males, we analyzed the sex differences in lymphocyte subsets, including CD4+ T cells, CD8+ T cells, B cells, and natural killer (NK) cells (Figure 2). The results showed that the percentages of B cells (9.1 [5.85-13.35] % vs. 11.2 [8.2-14.4] % in males and females, respectively, *P*<.001) and CD4+ T cells (38.6 [33.4-44.6] % vs. 42.9 [35.8-48.3] % in males and females, respectively, *P*<0.001) were remarkably higher in females than that in males. B cells play a pivotal an role in humoral immunity by differentiating to plasma cells under the stimulation of foreign antigens, and plasma cells can synthesize and secrete specific antibodies against virus infection [16]. Besides, CD4+ T cells are indispensable in helping the differentiation from B cells to plasma cells[17, 18]. The higher percentages of B cells and CD4+ T cells in females indicated that females had stronger humoral immunity than males during the infection and recovery of COVID-19. However, the CD8+ T cell percentage continuously increased within the 4 weeks after admission in males but maintained a relatively low level in females. The median percentage of CD8+ T cells was 29.7% and 23.6% in males and females, respectively, in the fourth week after admission. The CD8+ T cell percentage sharply decreased in males, while increased in females after the fourth week, indicating that the dynamic changes of cellular immunity were different between sexes. Moreover, NK cell percentage was higher in males than that in females throughout hospitalization (16.4 [11.3-23.3] % vs. 13.8 [8.6-18.4] % in males and females, respectively, *P*<0.001). These results suggested that males and females had different humoral and cellular immune responses in SARS-CoV-2 infection and recovery.

**Figure 2.**
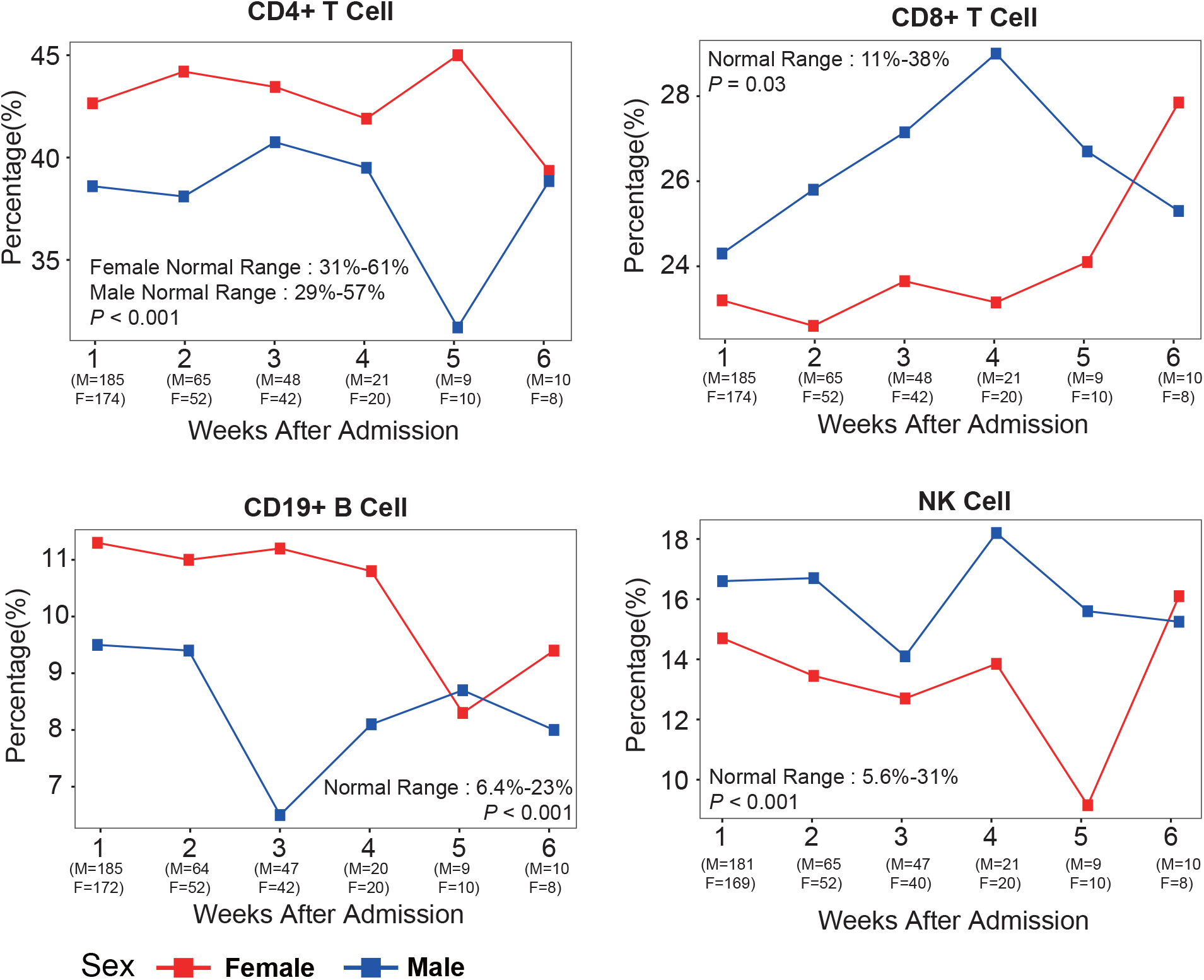
Comparison of lymphocyte subsets in peripheral blood between males and females. The x-axis displays the weeks after admission. F indicates the number of tests of females, and M indicates the number of tests of males. The y-axis displays the percentage of immune cells. Red line based on median is used to profile the variation tendency of the females, and blue line based on median is used to profile the variation tendency of the males.

### The response of SARS-CoV-2 specific IgG is more rapid in females than in males

As the observed stronger humoral immunity in females, we further compared the dynamic changes of total antibody, Spike protein (S)-, receptor binding domain (RBD)-, and nucleoprotein (N)-specific IgM and IgG levels during SARS-CoV-2 infection and recovery between males and females (Figure 3, Supplementary Figure S2). We observed similar dynamic trends of total IgM and IgG in males and females, except that the total IgG reached a relatively high level in the third week after symptom onset in females, while it took 4 weeks for the males to get the comparable antibody level. The N-specific IgG level showed no obvious differences between males and females. Notably, as one of the most important neutralizing antibodies against SARS-CoV-2 [19], the RBD-specific IgG level sharply increased within the first 4 weeks after onset in females. However, the RBD-specific IgG level increased much more slowly in males, and it took at least 7 weeks for males to reach a comparable level of the fourth weeks in females. The RBD-specific IgG levels were 11.3 AU/ML and 34.3 AU/ML in the fourth week in males and females, respectively. Moreover, the dynamic changes of the S-specific IgG level showed a similar trend with RBD-specific IgG. Our results indicated that although the S-and RBD-specific IgG levels continuously increased in males during the COVID-19 recovery, the response of these protective antibodies was much slower in males than that in females.

**Figure 3.**
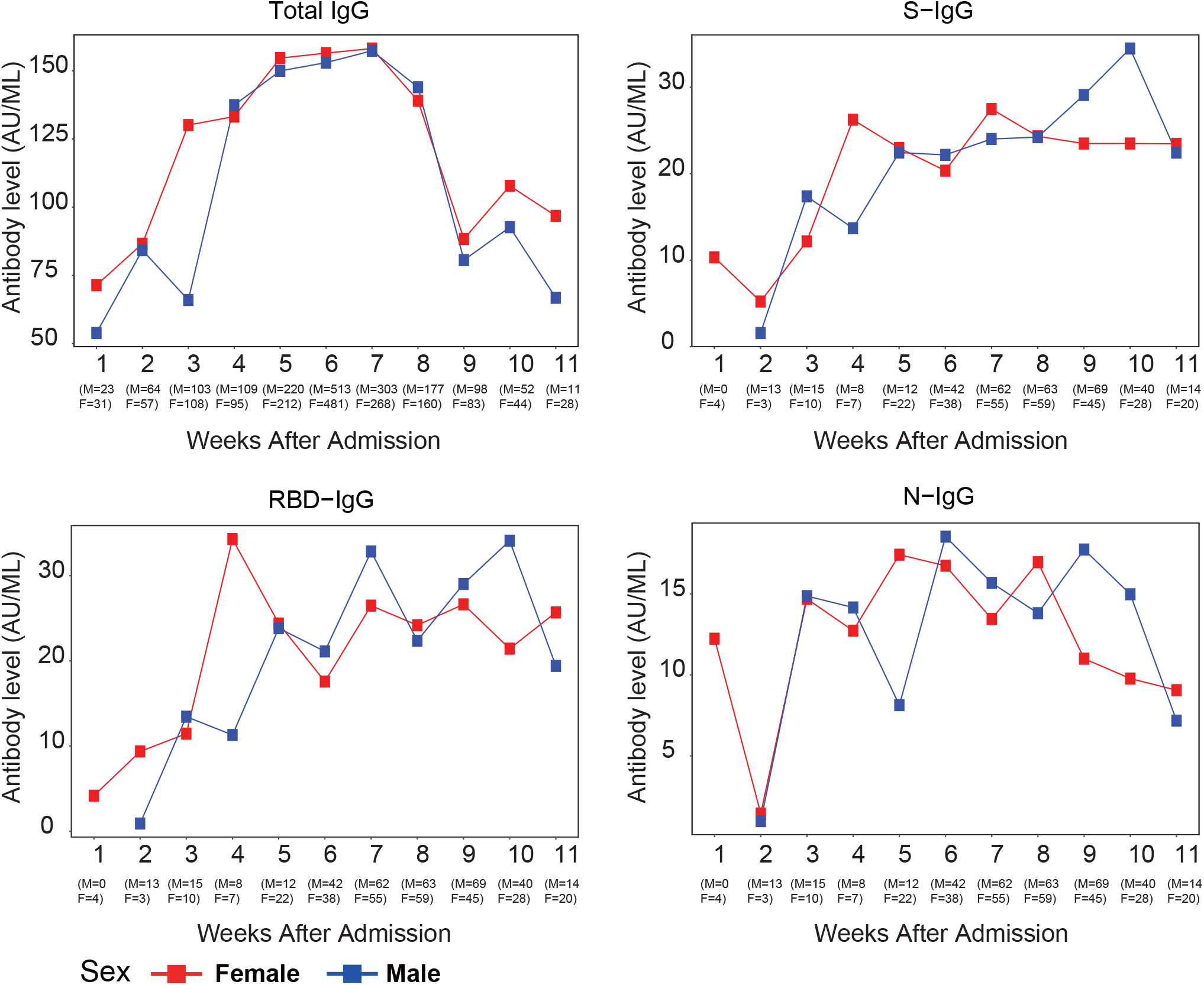
The dynamic changes of antibodies against SARS-CoV-2. The x-axis displays the weeks after admission. F indicates the number of tests of females, and M indicates the number of tests of males. The y-axis displays the level of IgG level. Red line based on median is used to profile the variation tendency of the females, and blue line based on median is used to profile the variation tendency of the males.

## Discussion

Although previous epidemiological studies reported that the mortality of males was significantly higher than females[20], the underlying cellular and molecular mechanisms have not been comprehensively analyzed due to insufficient sample size of test data. In this study, using a homogeneous data from a single-center, we described the clinical and laboratory characteristics of 3,057 COVID-19 patients (1,558 males and 1,499 females) and performed a comparative analysis for clinical outcomes and immunological responses between males and females. Our results showed that the mortality, ICU admission rate, and percentage of critical cases were approximately 2-fold higher in males than that in females. Sex is an independent prognostic risk factor for COVID-19. The concentration of inflammatory factors in peripheral blood was significantly higher in males, indicating a higher risk of the cytokine storm. Besides, the renal and hepatic abnormality induced by COVID-19 was more common in males during the hospitalization. The further immunological analysis revealed that females have stronger humoral immunity than males during the infection and recovery of COVID-19, and the response of protective antibodies was much slower in males than that in females.

In this decade, it is increasingly acknowledged that males and females differ in their immunological responses to foreign and self-antigens, including both innate and adaptive immune responses[21]. Studies showed that females have stronger immune responses against various diseases, such as HIV infection and cancer[22, 23]. Furthermore, for vaccination, females have greater responses and may experience greater efficacy than males [9]. The hormonal effect is one of the most important mechanisms that caused the sex differences in the immune response. For instance, estrogen has effects on various types of immune cells and is associated with lots of immune-related diseases[24]. Androgen response elements (AREs) and estrogen response elements (EREs) present in the promoters of immunity genes, indicating the sex steroids could have a regulatory function on immune response[25]. Notably, estrogenic compounds can reduce the influenza A virus replication in primary human nasal epithelial cells[26]. Treating female mice with estrogen receptor antagonists increased the mortality of SARS-CoV infection [10]. The evidence suggested that the estrogen treatment may help increase the immunity of males to defend against SARS-CoV-2 infection.

In addition, our results showed that the response of S-and RBD-specific antibodies were much slower in males than that in females after SARS-CoV-2 infection. The indolent antibody responses in males may lead to their rapid progression of the disease. This result indicated the importance of early medical intervention for males with COVID-19. Furthermore, immunotherapy such as convalescent plasma transfusion may enhance the immunity of males. Moreover, males have a higher risk of cytokine storm, and renal, hepatic abnormalities during the hospitalization. Closely monitoring of various indicators is necessary, especially for severe male patients.

## Conclusions

In this study, by analyzing the sex-based clinical and immunological differences in a large COVID-19 cohort, we found that the poor prognosis of male COVID-19 patients may due to the weak humoral immunity and indolent antibody responses during SARS-CoV-2 infection and recovery. Meanwhile, it is important for male patients to be early intervened and closely monitored.

## Perspectives and significance

Our results will provide important information for the epidemiology and precise medical intervention for COVID-19, and shed new light on the development of hormonal and immunological therapy for males.

## Data Availability

Data are available on request to the corresponding author

## Acknowledgements

We thank prof. Ying Xiao for her professional suggestions on drafting of the manuscript. We thank Ms. Tingting Zhang for her help with data analysis.

## Authors’ contributions

Qianghu Wang, Shukui Wang, and Xinyi Xia had full access to all of the data in the study and takes responsibility for the integrity of the data and the accuracy of the data analysis. Kening Li, Bin Huang, Yun Cai, Zhihua Wang contributed equally. Concept and design: Qianghu Wang, Shukui Wang, and Xinyi Xia. Experiments and data collection: Zhihua Wang. Data analysis and interpretation: Kening Li, Bin Huang, Yun Cai, Lu Li, Lingxiang Wu, Mengyan Zhu, Jie Li, Ziyu Wang, Min Wu, Wei Wu. Drafting of the manuscript: Kening Li, Bin Huang.

## Funding

This study was supported by the National Natural Science Foundation of China (Grant Nos. 81572893, 81972358, 81959113), Key Foundation of Wuhan Huoshenshan Hospital (Grant No. 2020[18]), Key Research & Development Program of Jiangsu Province (Grant Nos. BE2017733, BE2018713), Medical Innovation Project of Logistics Service (Grant No. 18JS005), Basic Research Program of Jiangsu Province (Grant No. BK20180036).

## Availability of data and materials

Data are available on request to the corresponding author

## Ethics approval and consent to participate

This study was approved by the Medical Ethical Committee of Wuhan Huoshenshan Hospital (HSSLL011), and the Ethical Committee of Nanjing Medical University (2020-511).

## Consent for publication

All authors consent to the publication of the manuscript in Biology of Sex Differences.

## Competing interests

We declare no competing interests.

## Notes

### Competing Interest Statement

The authors have declared no competing interest.

## Reference

1. Wu Z, McGoogan JM. Characteristics of and Important Lessons From the Coronavirus Disease 2019 (COVID-19) Outbreak in China: Summary of a Report of 72314 Cases From the Chinese Center for Disease Control and Prevention. JAMA. 2020; doi:10.1001/jama.2020.2648.

2. Grasselli G, Zangrillo A, Zanella A, Antonelli M, Cabrini L, Castelli A, et al. Baseline Characteristics and Outcomes of 1591 Patients Infected With SARS-CoV-2 Admitted to ICUs of the Lombardy Region, Italy. JAMA. 2020; doi:10.1001/jama.2020.5394.

3. Liu R, Han H, Liu F, Lv Z, Wu K, Liu Y, et al. Positive rate of RT-PCR detection of SARS-CoV-2 infection in 4880 cases from one hospital in Wuhan, China, from Jan to Feb 2020. Clin Chim Acta. 2020;505:172–5; doi:10.1016/j.cca.2020.03.009.

4. Guan WJ, Ni ZY, Hu Y, Liang WH, Ou CQ, He JX, et al. Clinical Characteristics of Coronavirus Disease 2019 in China. N Engl J Med. 2020;382:18:1708–20; doi:10.1056/NEJMoa2002032.

5. Zhu N, Zhang D, Wang W, Li X, Yang B, Song J, et al. A Novel Coronavirus from Patients with Pneumonia in China, 2019. N Engl J Med. 2020;382:8:727–33; doi:10.1056/NEJMoa2001017.

6. Jia N, Feng D, Fang LQ, Richardus JH, Han XN, Cao WC, et al. Case fatality of SARS in mainland China and associated risk factors. Trop Med Int Health. 2009; 14 Suppl 1:21-7; doi: 10.1111/j.1365-3156.2008.02147.x.

7. Karlberg J, Chong DS, Lai WY. Do men have a higher case fatality rate of severe acute respiratory syndrome than women do? Am J Epidemiol. 2004;159:3:229–31; doi:10.1093/aje/kwh056.

8. Xie J, Tong Z, Guan X, Du B, Qiu H. Clinical Characteristics of Patients Who Died of Coronavirus Disease 2019 in China. JAMA Netw Open. 2020; 3:4: e205619; doi:10.1001/jamanetworkopen.2020.5619.

9. Bischof E, Wolfe J, Klein SL. Clinical trials for COVID-19 should include sex as a variable. The Journal of Clinical Investigation. 2020.

10. Channappanavar R, Fett C, Mack M, Ten Eyck PP, Meyerholz DK, Perlman S. Sex-Based Differences in Susceptibility to Severe Acute Respiratory Syndrome Coronavirus Infection. J Immunol. 2017;198:10:4046–53; doi:10.4049/jimmunol.1601896.

11. Cai H. Sex difference and smoking predisposition in patients with COVID-19. Lancet Respir Med. 2020;8:4:e20; doi:10.1016/S2213-2600(20)30117-X.

12. Liu Y, Du X, Chen J, Jin Y, Peng L, Wang HHX, et al. Neutrophil-to-lymphocyte ratio as an independent risk factor for mortality in hospitalized patients with COVID-19. J Infect. 2020; doi: 10.1016/j.jinf.2020.04.002.

13. Qin C, Zhou L, Hu Z, Zhang S, Yang S, Tao Y, et al. Dysregulation of immune response in patients with COVID-19 in Wuhan, China. Clin Infect Dis. 2020; doi:10.1093/cid/ciaa248.

14. Mehta P, McAuley DF, Brown M, Sanchez E, Tattersall RS, Manson JJ, et al. COVID-19: consider cytokine storm syndromes and immunosuppression. Lancet. 2020;395:10229:1033–4; doi:10.1016/S0140-6736(20)30628-0.

15. Ni L, Ye F, Cheng ML, Feng Y, Deng YQ, Zhao H, et al. Detection of SARS-CoV-2-Specific Humoral and Cellular Immunity in COVID-19 Convalescent Individuals. Immunity. 2020; doi:10.1016/j.immuni.2020.04.023.

16. De Silva NS, Klein U. Dynamics of B cells in germinal centres. Nat Rev Immunol. 2015;15:3:137–48; doi:10.1038/nri3804.

17. Topham DJ, Tripp RA, Hamilton-Easton AM, Sarawar SR, Doherty PC. Quantitative analysis of the influenza virus-specific CD4+ T cell memory in the absence of B cells and Ig. J Immunol. 1996;157:7:2947–52; https://www.ncbi.nlm.nih.gov/pubmed/8816401.

18. Topham DJ, Doherty PC. Clearance of an influenza A virus by CD4+ T cells is inefficient in the absence of B cells. J Virol. 1998;72:1:882–5; https://www.ncbi.nlm.nih.gov/pubmed/9420305.

19. Tai W, He L, Zhang X, Pu J, Voronin D, Jiang S, et al. Characterization of the receptor-binding domain (RBD) of 2019 novel coronavirus: implication for development of RBD protein as a viral attachment inhibitor and vaccine. Cell Mol Immunol. 2020; doi:10.1038/s41423-020-0400-4.

20. Jin J-M, Bai P, He W, Wu F, Liu X-F, Han D-M, et al. Gender Differences in Patients With COVID-19: Focus on Severity and Mortality. Frontiers in Public Health. 2020;8; doi:10.3389/fpubh.2020.00152.

21. Klein SL, Flanagan KL. Sex differences in immune responses. Nat Rev Immunol. 2016;16:10:626–38; doi:10.1038/nri.2016.90.

22. Gandhi M, Bacchetti P, Miotti P, Quinn TC, Veronese F, Greenblatt RM. Does patient sex affect human immunodeficiency virus levels? Clinical Infectious Diseases. 2002;35:3:313–22; doi:Doi 10.1086/341249.

23. Li Y, Kang K, Krahn JM, Croutwater N, Lee K, Umbach DM, et al. A comprehensive genomic pan-cancer classification using The Cancer Genome Atlas gene expression data. BMC Genomics. 2017;18:1:508; doi:10.1186/s12864-017-3906-0.

24. Straub RH. The complex role of estrogens in inflammation. Endocr Rev. 2007;28:5:521–74; doi:10.1210/er.2007-0001.

25. Hannah MF, Bajic VB, Klein SL. Sex differences in the recognition of and innate antiviral responses to Seoul virus in Norway rats. Brain Behav Immun. 2008;22:4:503-16; doi:10.1016/j.bbi.2007.10.005.

26. Peretz J, Pekosz A, Lane AP, Klein SL. Estrogenic compounds reduce influenza A virus replication in primary human nasal epithelial cells derived from female, but not male, donors. Am J Physiol Lung Cell Mol Physiol. 2016;310:5:L415-25; doi:10.1152/ajplung.00398.2015.

